# Building National Patient Registries in Mexico: Insights from the MexOMICS Consortium

**DOI:** 10.1101/2023.09.11.23295377

**Authors:** Paula Reyes-Perez, Ana Laura Hernández-Ledesma, Talía V. Román-López, Brisa García-Vilchis, Diego Ramírez-González, Alejandra Lázaro-Figueroa, Domingo Martínez, Victor Flores-Ocampo, Ian M. Espinosa-Méndez, Miguel E. Rentería, Alejandra E. Ruíz-Contreras, Sarael Alcauter, Alejandra Medina-Rivera

**Affiliations:** Laboratorio Internacional de Investigación sobre el Genoma Humano, Universidad Nacional Autónoma de México, Campus Juriquilla, Blvd. Juriquilla 3001, 76230, Santiago de Querétaro, México; Departamento de Neurobiología Conductual y Cognitiva, Instituto de Neurobiología, Universidad Nacional Autónoma de México, Campus Juriquilla, Blvd. Juriquilla 3001, 76230, Santiago de Querétaro, México; Laboratorio de Neurogenómica Cognitiva, Unidad de Investigación de Psicobiología y Neurociencias, Coordinación de Psicobiología y Neurociencias, Facultad de Psicología, Universidad Nacional Autónoma de México. Av. Universidad 3004, Col. Universidad Nacional Autónoma de México, 04510, CdMx, México; Unidad de Genómica Avanzada, Langebio, Centro de Investigación y Estudios Avanzados del Instituto Politécnico Nacional, Libramiento Norte León Km 9.6, 36821, Irapuato, Guanajuato, México; Escuela Nacional de Estudios Superiores, Unidad Juriquilla, Universidad Nacional Autónoma de México, Juriquilla, 76230, Santiago de Querétaro, México; Mental Health and Neuroscience Program, QIMR Berghofer Medical Research Institute, 4006, Brisbane, Queensland, Australia; School of Biomedical Sciences, Faculty of Medicine, The University of Queensland, 4006, Brisbane, Queensland, Australia

**Keywords:** *Patient registries*, *genetics*, *twins*, *systemic lupus erythematosus*, *parkinson disease*

## Abstract

**OBJECTIVE:** To introduce MexOMICS, a Mexican Consortium focused on establishing electronic databases to collect, cross-reference, and share health-related and omics data on the Mexican population.

**METHODS:** The Mexican Twin Registry (TwinsMX), Mexican Lupus Registry (LupusRGMX) and the Mexican Parkinson’s Research Network (Mex-PD) were designed and implemented using Research Electronic Data Capture web-based application. Registries were compiled through voluntary participation and on-site engagement with medical specialists. In some instances, DNA samples and Magnetic Resonance Imaging images were also acquired.

**RESULTS:** Since 2019, the MexOMICS Consortium has successfully established three electronic-based registries: TwinsMX (n=2915), LupusRGMX (n=1761) and Mex-PD (n=750). In addition to sociodemographic, psychosocial, and clinical data, MexOMICS has collected samples for genetic determinations across the three registries. Cognitive function assessments, conducted using the Montreal Cognitive Assessment, have been administered to a subsample of 376 Mex-PD participants. Furthermore, a subset of 267 twins underwent measurements of structural, functional, and spectroscopy brain images; comparable evaluations are projected for LupusRGMX and Mex-PD.

**CONCLUSIONS:** The MexOMICS registries offer a valuable repository of information concerning the potential interplay of genetic and environmental factors in health conditions among the Mexican population.

## INTRODUCTION

### Brief history of patient registries

Patient registries are organized systems that collect, store and analyze information about individuals living with a particular health condition^1^. They are used for clinical research, recruitment into clinical trials, and improving patient care^2^. When coupled with genomic information, they provide further opportunities for genomic medicine and research^3^.

The National Leprosy Registry of Norway, established in 1856, was presumably the first national patient registry, implemented as a response of the high prevalence of leprosy at the time, and included a collaborative effort between health officers and ministers of the church, which resulted in the policy of isolation that ultimately led to the disappearance of leprosy in Norway^4^.

Since then, several registries have been set around the globe with various aims. As expected, the two leading causes of death, cancer and heart diseases, have led the efforts through the past century. Nowadays, there are at least 343 cancer registries in 65 countries^5^, while 155 Cardiac disease registries in 49 countries have enrolled 73.10 million patients^6^. One of the greatest examples of this is the US National Cardiovascular Data Registry (NCDR), which comprises 10 registries and has more than 60 million records now entering the qualification of Big Data^7^. In Latin America, population-based cancer registries, which arguably are the most extensively documented among patient registries, cover 20% of the population, and only 7% of these are considered to contain high-quality information^8^.

Although usually patient registries aim to estimate the prevalence of a disease or trait within a population, and establish what demographics and environmental factors affect risk, they don’t always aim to estimate how much a trait is explained by genetic or environmental factors. Historically, twins’ registries have been frequently used to improve our comprehension of human health and the genetic and environmental basis of complex diseases^9^. The classical twin design consists of acquiring phenotypic data of monozygotic (MZ) and dizygotic (DZ) twins and measuring how correlated a trait is within pairs of both types. The heritability of a trait, the percentage of variance explained by genetic factors, can be estimated through the difference between the correlation of MZ twins and DZ twins. The rest of the variance in that trait could be explained by environmental factors. Other research designs might also acquire data from other family members of the twin participants, the so-called extended twin design, or acquire data of MZ twins adopted to different households. However, twin studies are not just limited to estimating the heritability of a trait, they can also estimate how two different traits have genetic and environmental factors in common explaining their variance. They can also aid in establishing causal relationships between two associated traits, for example, MZ twins discordant in cannabis consumption could help to determine whether cannabis does cause lower cognitive functioning or not. Thus, the information generated through twin registries has allowed us to deepen the genotype-environment interactions, how they shape the mechanisms that lead us to our individual differences and the influence of these mechanisms in health and disease conditions^9^. Furthermore, Latin America has allegedly the smallest sample size for twin registries, of around 5000 individuals, Brazil being the country with most registries (n=4826), followed by Mexico^9^.

Thus the often mandatory implementation of patient records and twin registries across some countries has promoted relevant progress^10^. Nevertheless, still a considerable fraction of the world’s population remains uncovered, particularly in African and Asian countries^11^, due to the struggle to collect and integrate data to establish and maintain national registries^12^.

### The context of Mexican health records systems

Although healthcare in Mexico has certainly improved its coverage and scope over the last two decades^13^, according to the 2020 census, about 33 million people in the country remain with no health services insurance^14^ and 48.49% have no effective access to health services^13^.

Considering that leading causes of death in Mexico include heart disease, diabetes and malignant tumors^15^ initiatives such as the Mexican Registry of Atrial Fibrillation (ReMeFa)^16^, the Mexican Registry of Pediatric Cardiac Surgery^17^ and the National Registry of Cardiac rehabilitation programs (RENAPREC), have been recently established. But it seems that in some cases efforts must be reinforced; for instance, the National Heart Failure Registry Program^18^ was initiated in 2002 with the sole purpose of having a general characterization of patients with this condition, once the aim was achieved in 2004, the project was no longer pursued.

In recent years, valuable efforts have been made to establish registries in Mexico for patients living with conditions such as osteoarthritis^19^, diabetes^20^, atrial fibrillation^21^. Although a federal norm (NOM-024-SSA3-2012, issued in 2012) was established to regulate the Electronic Health Record Information Systems^22^, the diversity of electronic clinical records and database storage systems greatly hinder data sharing and interaction among health institutions^23^. In addition, these registries are mainly survey-based and lack biobanking or genetic analyses, unlike their homologs in the European population, which limits their scope^24–26^.

The MexOMICS Consortium was created with the aim to design and implement an infrastructure that allows the consolidation of electronic-based databases with the ability to collect, compare, cross-reference, and share valuable information about Mexican people with different healthcare related conditions. The consortium is currently focused on three traits in the admixed Mexican population: a) twins and members of multiple births (*TwinsMX*)^27^, b) people with Systemic Lupus Erythematosus (*LupusRGMX)*^28^ and c) patients with Parkinson Disease (*MEX-PD)* ^29^. To our knowledge, there is a lack of other nationwide registries that include genetic data, and publicly available information characterizing Mexican people with these traits In addition to the valuable clinical and sociodemographic information that the developed databases could provide, these include genetic profiling and biobanking with the aim to expand our knowledge of the influence of genetic-environmental interactions on the phenotypic characteristics of the Mexican population and their potential implications on health and disease. In the present work, we will discuss the design, implementation and operational issues, current state and future directions of these three registries.

## METHODS

### Ethics and data protection

All three registries were approved by the Ethics on Research Committee of the Institute of Neurobiology at Universidad Nacional Autónoma de México (UNAM). TwinsMX was the first registry, established in 2019; followed by LupusRGMX and MEX-PD in 2021. Information is anonymized and stored at the National Laboratory of Advanced Scientific Visualization at UNAM. Participants provided informed consent prior to registration, and a privacy statement was given to them, in accordance with the Federal Law on the Protection of Personal Data Held by Private Parties.

### Platform and surveys design and implementation

Each registry follows specific guidelines in order to obtain necessary data. For such purposes, surveys are available on web-based application Research Electronic Data Capture (REDCap), which allows secure capture and storage of data^30,31^. Instruments integrated in the three registries are summarized in **Table 1** to provide a general picture of the type of information that is being collected.

**Table 1.**
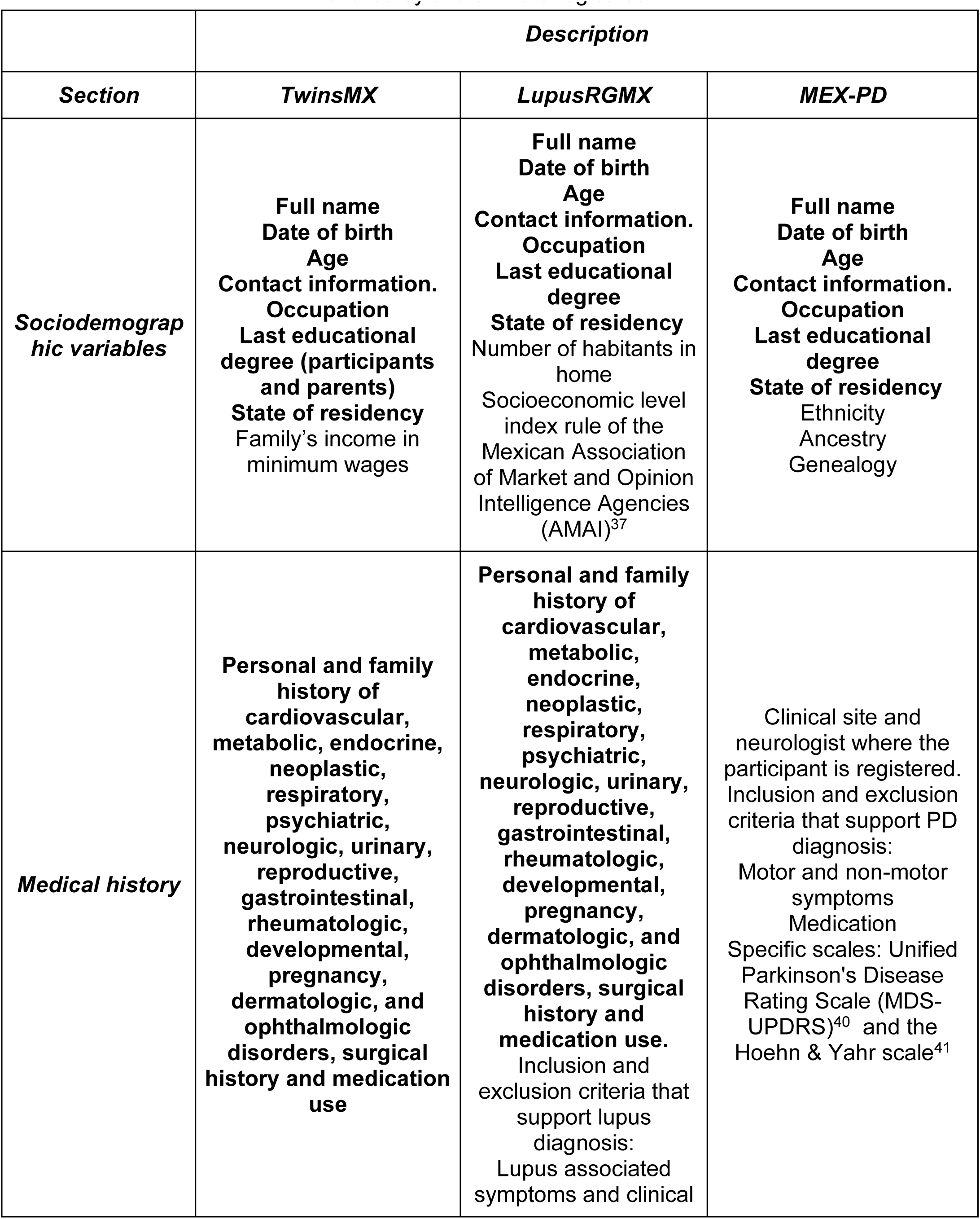
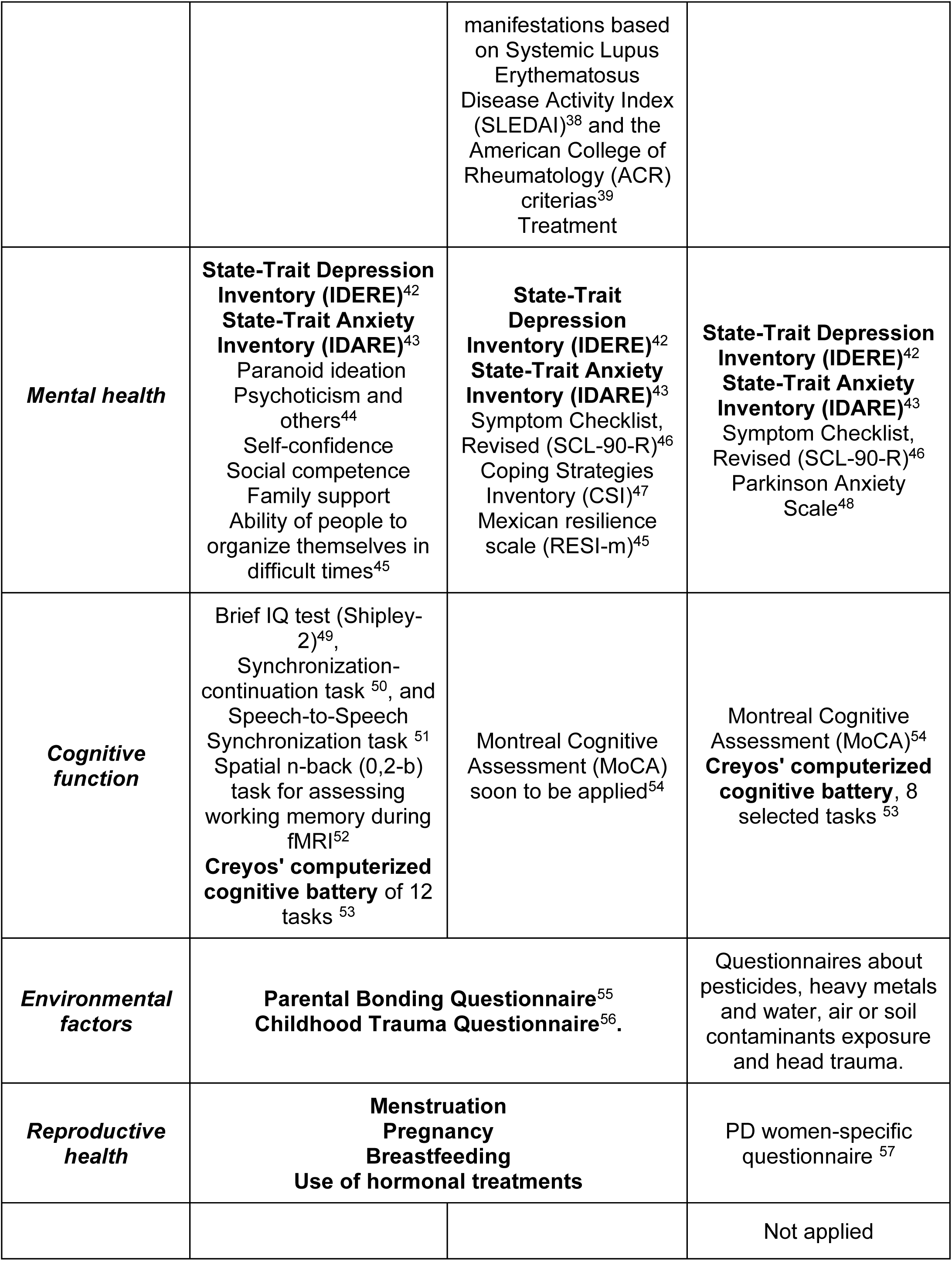
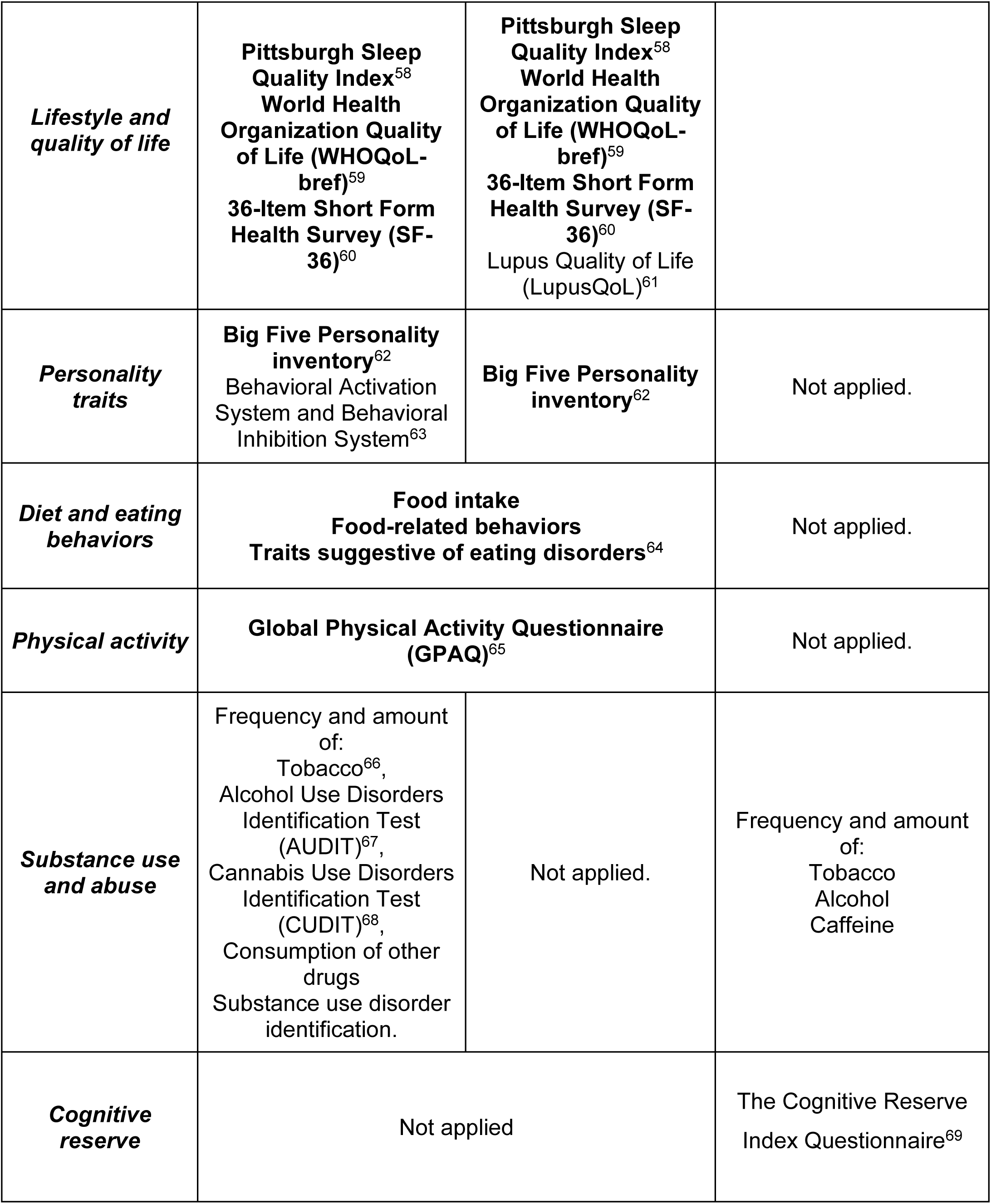
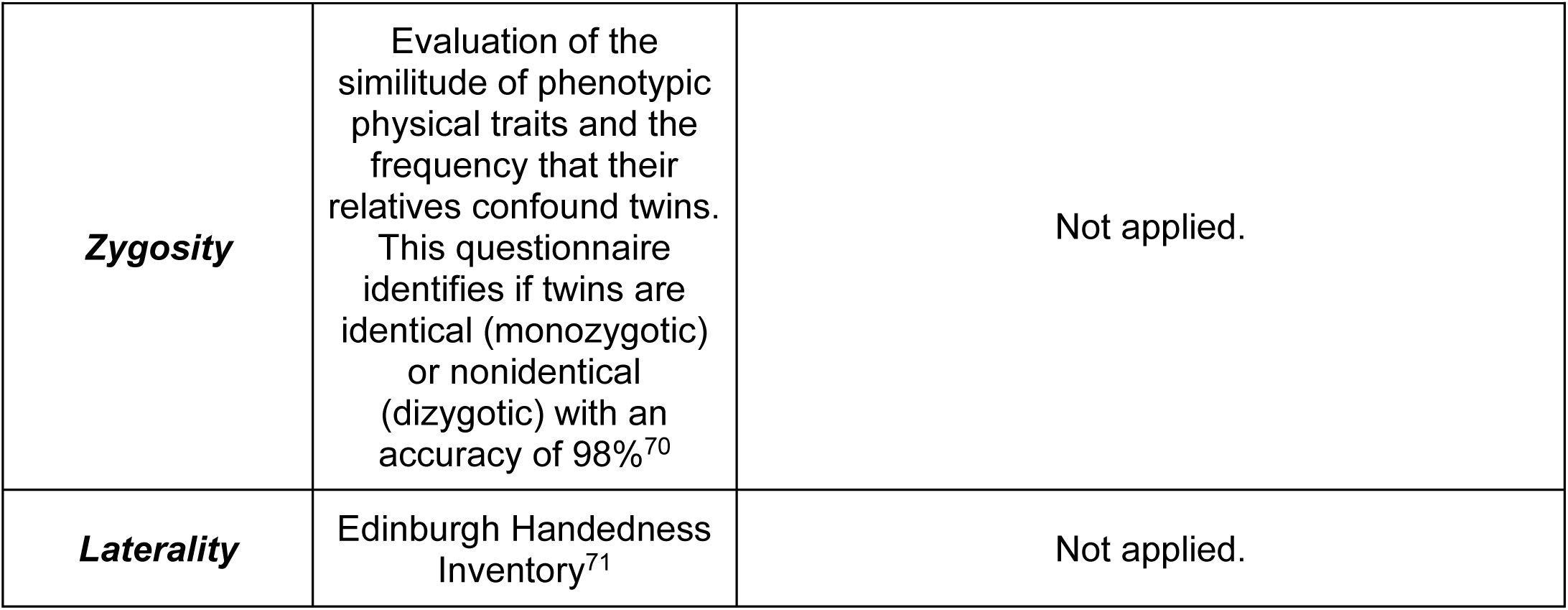
Instruments employed and information required in TwinsMX, LupusRGMX and MEX-PD registries. In bold letters are presented the variables and instruments shared by two or more registries.

In general, all three projects share an initial structure, with questionnaires focused on identifying socio demographic traits, characterizing medical history and mental health. The shared technology information infrastructure of the registries has allowed us to design, implement and validate surveys in one of the registries to further establish it in the other registries. Beside these shared questionnaires, each registry includes surveys focused on bringing light on specific traits associated with the studied population, such as zygosity in TwinsMX or cognitive impairment in MEX-PD.

Although the selection of each instrument responds to particular research questions one major guideline is the ability to produce harmonized measures to link and compare the data across registries and their counterparts in other populations ^32^. For instance, TwinsMX is built to be comparable to the Australian ^33^ and United Kingdom ^34^ Twins Registries’, while MEX-PD is akin to the newly established Australian Parkinson’s Genetics Study^35^ and the Latin American Research Consortium on the Genetics of Parkinson’s Disease^36^.

### Project identity

The term *-omics* has been widely used in recent years to address specific fields in biological sciences research, such as the study of the genome (*genomics)* or the study of the metabolome (*metabolomics*)^72^. MexOMICS reflects our aim to study health related conditions in the Mexican population from a multidisciplinary approach; evaluating not only the clinical data, but also integrating the role of genetic and environmental factors. As for the logo, “MexOMICS” name is accompanied by four entangled DNA strands each representing the conjunction of projects (TwinsMX, LupusRGMX and Mex-PD) in a Mexican styled design that resembles the *ollin* (movement) prehispanic symbol.

For the Mexican Twin Registry, Registro Mexicano de Gemelos in Spanish, the acronym TwinsMX was selected; “Twins” to highlight the multiple births and “MX” for Mexican. The logo is centered in this acronym and includes “Registro Mexicano de Gemelos” in the bottom part. The “I” and “X” letters include two dots on the top, resembling a pair of heads, representing the pairs of twins (Figure 1B). Color palette includes bright green and a shade of pink known as Mexican pink in our culture; both colors are widely used on traditional artisanal crafts and therefore represent the vibrancy of our culture, as well as the spirit, charisma, liveliness, and vitality of our people. TwinsMX’s website (https://twinsmxofficial.unam.mx/) includes the “Sign up” section, along with an “About us” section focused on the academic profiles of the TwinsMX team members, frequently asked questions, information about the projects well as interactive section that allow the people to see how is the registry advancing through time. It also includes a “Blog” section focused on sharing science-based information of interest in a non-technical language. TwinsMX was launched in May 2019 and represents the first platform implemented by our consortium.

**Figure 1.**
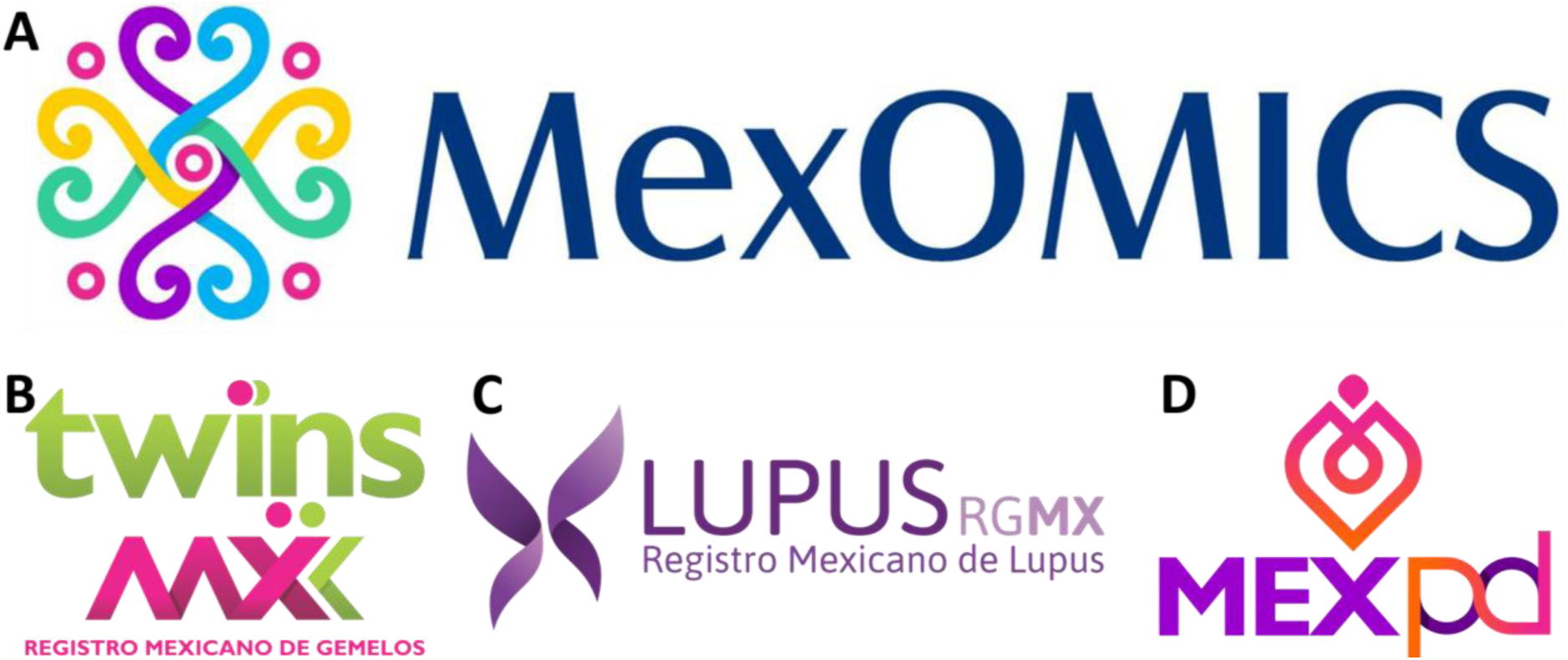
Logos designed for A) MexOMICS, B) TwinsMX, C) LupusRGMX and D) MEX-PD.

For the Mexican Lupus Registry’s acronym, the word Lupus was selected as the central element; followed by the letters “RG”, from ReGistry and “MX”, from MeXican. The registry’s logo includes the acronym along with “Registro Mexicano de Lupus” in the bottom part, and integrates a butterfly, which resembles the malar rash, one of the most common manifestations of SLE, along with a meaning of hope; the purple color palette was selected because of its wide utilization in awareness campaigns as it symbolizes a middle point between the intensity and motivation of the red and the calmness and stability of the blue (Figure 1B). The website (https://lupusrgmx.liigh.unam.mx/) was created considering the same color palette and it includes a “who are we” section which includes academic profiles of the research team members, a “Communication” part with the structure of an informative blog, a “Lives” section that resumes the facebook lives hosted by members of the project, a “Contact” section to provide a communicative platform with the team, and the “Sign up” button to access to the registry. LupusRGMX was successfully launched in May 2021, was socialized through a live session via facebook, and have worked since then using these platforms.

The Mexican Parkinson’s Research Network, Red Mexicana de Investigación en Parkinson in Spanish, is represented with the acronym “MEX-PD” which includes “MEX” for Mexican and “PD” for Parkinson Disease (Figure 1C). The logo includes a tulip silhouette, a flower that has been broadly used to raise awareness about Parkinson disease since a horticulturist living with PD developed a new variant of tulip and named it after Dr. James Parkinson, the first physician to describe and document the disease. MEX-PD website (https://mexpd.liigh.unam.mx/) includes a “Frequent asked questions” section, an “About us” section that includes the academic information about the team and a “Pre-registry” option. MEX-PD was launched in July 2021.

### Volunteers’ recruitment

As one of the main objectives of the registries is to include as many Mexicans as possible across the country, it became evident that a decentralized approach was needed. Given the varied nature of each registry, specific procedures for participants recruitment have been followed: for TwinsMX the enrollment is made through social media campaigns and on-site events invitations. Given the complexity of PD diagnosis the recruitment in MEX-PD requires the clinical expertise of neurologists; a team of neurologists of different sites across México has been assembled and volunteers are being identified and registered through their practice. For LupusRGMX the recruitment relies on a hybrid strategy: A team of rheumatologists is actively registering patients in their practice, and through social media and on site events where volunteers are invited to self-register.

## RESULTS

### Progress at a glance

As of June 2023, our registries included 2915 volunteers for TwinsMX (April 28th, 2023), 1761 participants for LupusRGMX (June 21st, 2023) and 750 for MEX-PD (June 29th, 2023) (Table 2); additionally, for LupusRGMX 153 participants were registered as controls, whereas MEX-PD has 397 volunteers registered as non-affected controls. The female participation represents the 71.9% of TwinsMX, the 94.3% in LupusRGMX and the 59.2% in MEX-PD, whereas 28.1%, 6.7% and 40.8% of male participation was respectively observed in each registry.

**Table 2.** Sociodemographic characteristics of volunteers across registries.

| Information | TwinsMX<br>(n=2915*) | LupusRGM<br>X (n=1761) | MEX-PD<br>(n=750) |
| --- | --- | --- | --- |
| Gender (n,%)<br>*Female*<br>*Male* | 2096 (71.9%)<br>819 (28.1%) | 1662<br>(94.3%)<br>99 (6.7%) | 444<br>(59.2%)<br>306<br>(40.8%) |
| Age at enrollment (mean,SD) | 29.2±11.2 | 37.1±10.9 | 62.17±10.6 |
| Time with the diagnosis (mean,<br>SD) | NA | 8.5±7.8 | NA |
| Age at diagnosis (mean,SD) | NA | 28.3±10.9 | NA |
| Genetic samples obtained (n,%) | 232 (7.9%)** | 108 (6.1%) | 495 (66%) |
| MRI sampled (n) | 267 (9.1%) | To start in<br>fall 2023 | To start in<br>fall 2023 |
| Cognitive function evaluations | 267 (9.1%)** | To start in<br>fall 2023 | 376<br>(50.1%) |

\* Individual subjects
\*\* Subjects in MRI study

Since 2020 TwinsMX includes participation of pediatric volunteers, always with the consent of a parent or legal tutor, this is reflected in the age distribution of the cohort, which covers a range from 0 to 80 y/o, with the majority among 20 and 40 y/o (Figure 2A1), and a mean age of 29.2±11.2 y/o at the moment of enrollment. Distribution among monozygotic and dizygotic participants is shown in Fig XX, and a similar pattern can be observed in both groups.

**Figure 2.**
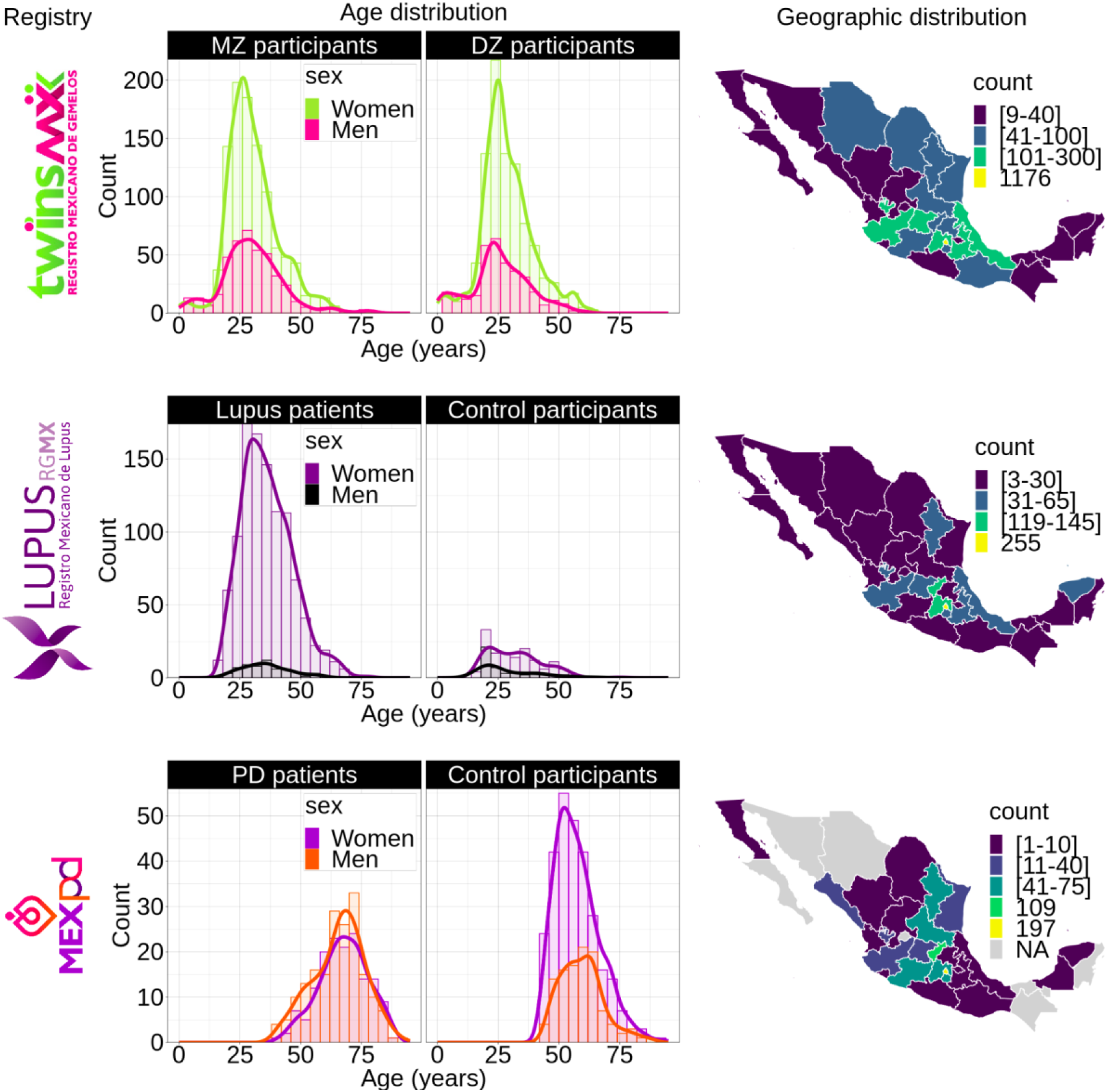
Geographic and age distribution of volunteers for TwinsMX (n=2915), LupusRGMX (n=1761) and E=MEX-PD (n=774). Age distribution for TwinsMX (upper-left), LupusRGMX (middle-left) and MEX-PD (bottom-left). Geographic distribution for TwinsMX (upper-right), LupusRGMX (middle-right) and MEX-PD (bottom-right).

In the case of LupusRGMX only adults (people over 18 y/o) can participate, age range in the registry is between 18 to 80 y/o(Figure 2B1), with the majority of the cohort between 20 and 45 y/o, with a mean age of diagnosis of 28.3±10.9 y/o. Healthy controls age distribution is also exhibited, with a range from 10 to 60 y/o, and the majority of volunteers being in their early 20s.

For MEX-PD the age of participants ranges from 40 to 90 y/os (Figure 2C1), this is due to the selection criteria that requires a minimum age of 40y/o, with the majority being between 60 and 80 y/o and a mean age of 62.17±10.6 y/o. Control participants exhibited a similar range, but with many of the volunteers among 40 and 60 y/o.

As mentioned, genetic analyses and other clinical measurements can help enrich the information collected through the registries, giving insights into the biological conditions of our participants. In this line, our team has been recovering DNA samples from volunteers of the three projects. To date, 109 samples have been recovered to perform whole genome sequences in LupusRGMX, of which 62 have been successfully sequenced; simultaneously 50 samples have been obtained to perform single cell RNA-seq. In the case of MEX-PD 495 samples have been obtained to perform genotyping. TwinsMX has a total of 232 available samples, of which 94 have already been genotyped, 82 are ready for genotyping, and the rest will be processed and genotyped in the next months.

As the nervous system is highly compromised in Parkinson’s disease, cognitive function has also been evaluated by the Montreal Cognitive Assessment (MoCA) to 126 controls and 250 patients. Due to the high prevalence and severity of neurological related manifestations in SLE (also known as neurolupus) LupusRGMX will apply 90 MoCA evaluations by 2024. Additionally, structural, functional and spectroscopy brain images have been acquired through MRI sessions of a subsample of 267 TwinsMX volunteers. MEX-PD and LupusRGMX will perform at least 90 MRI evaluations each by 2024.

Regarding geographical coverage of the three projects, we have an important participation of volunteers from the center of México, this is probably due to the physical location of the leading laboratories (Mexico City and Queretaro). In TwinsMX (Figure 2A2) the majority of registers are concentrated in México City (n=1176), México (n=295), Veracruz (n=149), Jalisco (n=111), Puebla (n=105), and Guanajuato (n=101), with less participation among people from the north-western and south regions. For LupusRGMX (Figure 2B2) we can observe a higher participation among people living in México City (n=255), México (n=145) and Querétaro (n=119); whereas the west and northern west regions are the less represented. In the case of MEX-PD (Figure 2C2) most of the registries come from México City (n=197), Querétaro (n=109), Morelos (n= 75), Michoacán (n=72), San Luis Potosí (n=71), México (n=62), and Nuevo León (n=42); the states with no volunteers registered are shown in light gray.

### Challenges and limitations

The implementation of a National Registry always faces challenges, especially at the beginning. For instance, TwinsMX relies completely on self-registration of volunteers that are reached by our invitations through social media or events, since there is no database available that allows us to identify and contact potential participants, which limits the scope of the project. Besides, it is necessary that all the siblings of a multiple birth complete the surveys; but in most cases, only one of the twins is interested or motivated to finish the questionnaires, which leads to incomplete data. Another limitation is that in Spanish there is a word specifically used for fraternal twins “*cuate”* (/’kwate/), thus often they do not identify themselves as twins, making them think that they are excluded from the registry. Stronger and more specific media campaigns are currently trying to address this issue.

LupusRGMX mainly works on a voluntary self-registration basis of people reached by our communication campaigns, by far, the main obstacle we have faced has been reaching Mexican individuals with SLE with the interest and disposition to start and complete the surveys: this is reflected on the fact that from the 1761 started registries, only 1271 have completed the clinical questionnaire. Different strategies, such as spacing out survey’s invitations, and motivation for participants through dynamics, such as giveaways, have been undertaken in order to increase survey completion. In addition, since the beginning, LupusRGMX, has tried to approach rheumatologists through the Mexican College of Rheumatology, in order to establish collaborations in which the rheumatologist gives a formal clinical evaluation at the beginning of the registry and help their patients through the initial clinical manifestations’ questionnaires, improving the reliability of our data. To date, only 147 participants have been registered through this option. With the knowledge that self-reported data can be threatened by self-reporting bias and thus considered as unreliable ^73^, strategies are being undertaken to assess them; we are currently working on a comparison of the outcomes from self-reported instruments with those provided by patients’ medical practitioners to evaluate reliability.

Similarly, as MEX-PD patients must be registered through a Neurologist with expertise to perform a clinical evaluation, reaching people with access to this level of medical care has been challenging. The shortage of Neurologists with valid certification in Mexico ^74^ along with their substantial workloads, driven by the necessity to attend to a large number of patients, limits the scope of the registry. Nonetheless, 774 registries have been recorded so far. Furthermore, as aforementioned, Neurologists in Mexico usually have a heavy workload, making the survey’s completion a daunting task. To ease this task, a team of trained assistants help patients complete particular surveys (i.e. environmental exposure), moreover, we are currently working on transferring suitable instruments to be self-completed by participants.

Likewise, a shared challenge between the registries is the need to acquire data from more diverse individuals within Mexico. Reviewing the current progress of the cohorts, it became evident that the majority of registries are representative of the central states of México and from specific regions where we have collaborating associations. For such a reason, we are working on specifically targeting the underrepresented regions of the country, taking advantage of our online tools and the use of social media and the establishment of new collaborations. Collection of biological samples through buccal swabs for DNA obtention is being performed in multiple sites for TwinsMX and MEX-PD, which will allow representation of diverse genetic backgrounds. However, Magnetic Resonance Images acquisition, performed in suitable TwinsMX participants, and blood samples for LupusRGMX, demand on-site presence of participants and are only carried out in Queretaro, limiting the participation of people from northern or southern states in the country. The complexity of this technique and its required infrastructure prevents its mobility to other Mexican states.

Finally, it is relevant to mention that due to that National registries are not common in the Mexican population, constant invitation in traditional and social media is required to continuously boost participation. As of 2020, 72.0% aged six or older Mexico residents are internet users ^75^, therefore the recruitment through social media has shown to be a very effective tool with similar models successfully implemented in other registries ^76,77^. To date, our registries have implemented an integrative approach through social media, turning them into reliable platforms for the exposition and discussion of topics associated with the interest of the registry’s community.

### Sharing is caring: community building

As a consortium, we strongly believe that the participants in the three registries are not subjects of study, but active citizens with a major role in biomedical research. As such, it is imperative to observe informed consents, maintain transparent procedures, disclose progress reports, and open communication channels where participants can provide feedback.

As twins, Lupus and Parkinson communities in Mexico were already active, one of the first steps in building the three registries was approaching community leaders, civil organizations, and foundations, which have been a key component in reaching participants and gaining credibility in their communities. TwinsMX, the first of this consortium, received support from the Multiple Birth Association (Asociación de Nacimientos Múltiples, A.C.), but now it holds a significant community by its own, with an important presence in social media with over 3.2K followers in facebook (https://www.facebook.com/TwinsMXofficial) and 748 on instagram, (https://www.instagram.com/twinsmxofficial). To date, TwinsMX’s team works constantly in the generation of informative content (https://twinsmxofficial.unam.mx/blog/) and their participation in community activities.

Furthermore, LupusRGMX receives direct support and active participation of representatives of the biggest communities of people with Lupus in the country: LUPUS MX (https://www.facebook.com/LUPUSMXOficial), Fundación Proayuda Lupus Morelos A.C. (https://www.facebook.com/LUPUSMORELOS), the Centro de Estudios Transdisciplinarios Athié-Calleja por los Derechos de las Personas con Lupus A.C. (https://www.facebook.com/Cetlu), and Despertar de la Mariposa A.C. (https://www.facebook.com/DespertarLupus); who have reviewed the surveys, and expressed their opinions and needs. We have also reached rheumatologists and the Colegio Mexicano de Reumatología, in order to establish collaborations that would help to provide a more integrated evaluation of SLE in our population. The formation of this community has helped us identify necessities and areas of opportunity for clinicians and researchers in order to generate knowledge with future applications and impact on the life of people with lupus. Through the contact of experts on some topics of interest such as pediatric SLE, grief associated with a chronic disease diagnosis, and reproductive health we have established monthly open talks through Facebook Live sessions (https://www.facebook.com/lupusrgmx). Beside these talks, the team of LupusRGMX has been working on providing another sources of reliable information (https://lupusrgmx.liigh.unam.mx/comunicacion.html) and actively participate in activities for lupus acknowledgment in México.

Similarly, MEX-PD has seeked support from the Red Mexicana de Asociaciones de Parkinson (https://www.facebook.com/RedMXdeAsociacionesdeParkinson), and different communities of people with Parkinson’s and their close ones, including Parkinson Laredo (https://www.facebook.com/groups/768321520680203) and Asociación Mexiquense de Parkinson IAP (https://www.facebook.com/parkinsonmexiquense).

### Perspectives

As a general perspective, TwinsMX, LupusRGMX and MEX-PD will allow us to characterize the genetic and environmental contribution over different traits and diseases with data directly collected in the Mexican population, which is heavily underrepresented in epidemiological and genetic studies. This characterization will provide us medically relevant information that in the future can be useful for better and more accurate diagnosis, treatments, and therapies.

In particular, for TwinsMX we expect to start to collect DNA through saliva samples, not only in twin individuals but also to their direct relatives (i.e. parents and not-twins siblings) in order to build a biobank with valuable data about genetic variants that make Mexicans more prone to develop relevant medical conditions. Also, in the future it is expected to acquire data with electroencephalography (EEG) and to be able to establish an association with cognitive performance. Then, taking together the EEG data, the data from the TwinsMX online registry and the MRI images, we will be able to establish associations between different traits and structural and functional brain properties.

Meanwhile, LupusRGMX has started applying behavioral evaluation of decision making, focused on the choice between social and material incentives and the temporary discount rate of such incentives. We are currently evaluating if regular glucocorticoid consumption is associated with delayed reaction times in decision-making. Additionally, magnetic resonance images and cognitive function evaluations will be acquired from ninety participants to explore the underlying functional and structural brain biomarkers. Furthermore, as a result of community feedback, LupusRGMX will expand surveys to collect data about reproductive health, allergies, and soon, we will be able to analyze and integrate information recovered from the registry with biomarkers and clinical data.

Finally, MEX-PD is currently performing a control participants recruitment campaign, for which we are working in finding collaborators in as many sites as possible and opening self-completed surveys. Also, as aforementioned, in a short time, we will begin data acquisition for structural and functional MRI, cognition and mental health.

## CONCLUSION

Patient registries are fundamental tools in public health research, providing comprehensive population-based data on diseases’ etiology and progression. The establishment of registries in Mexico, particularly those initiated by the Universidad Nacional Autónoma de México through the MexOMICS Consortium, has enriched the research landscape with epidemiological and genetic information. These registries enable researchers to explore the complex interplay between genetic factors and environmental influences in traits in the general population by studying twin participants, but also those in complex diseases like systemic lupus erythematosus, and Parkinson’s disease. However, limitations, such as inconsistent data entry and patient participation, together with potential biases due to voluntary participation must be addressed to ensure that the creation of these databases can serve as robust foundations for subsequent studies and investigations. The MexOMICS Consortium’s registries represent a valuable resource that will help to approach questions associated with health and disease in Mexican people. Future directions involve refinement and expansion, optimizing experimental design, data collection methodologies, and fostering interdisciplinary collaboration.

## ACKNOWLEDGMENTS

This work received support from Luis Aguilar, Alejandro León, and Jair García of the Laboratorio Nacional de Visualización Científica Avanzada. We also thank Cesar Arturo Dominguez Frausto, Erick H. Pasaye Alcaraz, Leopoldo González Santos, Carina Uribe Díaz, and Alejandra Castillo Carbajal for their technical support. We thank Mauricio Guzman for designing all logos.

Authors would like to express their special acknowledgment to Fundación Proayuda Lupus Morelos A.C., Lupus MX, El despertar de la Mariposa A.C., Centro de Estudios Transdisciplinarios Athié-Calleja por los Derechos de las Personas con Lupus A.C. (Cetlu), Lupus Tuxtla, Lupus en Yucatán, Asociación de Lupus y AIJ Caminando Juntos A.C., Aprendiendo a Vivir con Lupus y Fibromialgia A.C. and Asociación de Nacimientos Múltiples, A.C. for their invaluable support.

## DATA AVAILABILITY

All data is available upon request to authors, expression of interest can be done through this platform (https://redcap.link/qr4lspyl).

